# A microbiota-directed complementary food intervention in 12-18-month-old Bangladeshi children improves linear growth

**DOI:** 10.1101/2024.04.29.24306564

**Authors:** Ishita Mostafa, Matthew C. Hibberd, Steven J. Hartman, Md Hasan Hafizur Rahman, Mustafa Mahfuz, S. M. Tafsir Hasan, Per Ashorn, Michael J. Barratt, Tahmeed Ahmed, Jeffrey I. Gordon

## Abstract

**Background:** Globally, stunting affects ∼150 million children under five, while wasting affects nearly 50 million. Current interventions have had limited effectiveness in ameliorating long-term sequelae of undernutrition including stunting, cognitive deficits and immune dysfunction. Disrupted development of the gut microbiota has been linked to the pathogenesis of undernutrition, providing potentially new treatment approaches.

**Methods:** 124 Bangladeshi children with moderate acute malnutrition (MAM) enrolled (at 12-18 months) in a previously reported 3-month RCT of a microbiota-directed complementary food (MDCF-2) were followed for two years. Weight and length were monitored by anthropometry, the abundances of bacterial strains were assessed by quantifying metagenome-assembled genomes (MAGs) in serially collected fecal samples and levels of growth-associated proteins were measured in plasma.

**Findings:** Children who had received MDCF-2 were significantly less stunted during follow-up than those who received a standard ready-to-use supplementary food (RUSF) [linear mixed-effects model, β_treatment group x study week_ (95% CI) = 0.002 (0.001, 0.003); *P*=0.004]. They also had elevated fecal abundances of *Agathobacter faecis*, *Blautia massiliensis*, *Lachnospira* and *Dialister*, plus increased levels of a group of 37 plasma proteins (linear model; FDR-adjusted *P*<0.1), including IGF-1, neurotrophin receptor NTRK2 and multiple proteins linked to musculoskeletal and CNS development, that persisted for 6-months post-intervention.

**Interpretation:** MDCF-2 treatment of Bangladeshi children with MAM, which produced significant improvements in wasting during intervention, also reduced stunting during follow-up. These results suggest that the effectiveness of supplementary foods for undernutrition may be improved by including ingredients that sponsor healthy microbiota-host co-development.

**Funding:** This work was supported by the BMGF (Grants OPP1134649/INV-000247). ClinicalTrials.gov identifier: NCT04015999

## Introduction

Childhood undernutrition is a public health priority manifested by impaired ponderal and linear growth (wasting and stunting), immune and metabolic dysfunction, altered central nervous system (CNS) development as well as other physiologic abnormalities^1^. Undernutrition is typically classified using anthropometric assessments. It is estimated that globally, ∼50 million children under the age of five suffer from acute malnutrition (wasting); those with moderate acute malnutrition (MAM) have weight-for-length Z scores (WLZ) that are 2-3 standard deviations below the median (WLZ −2 to −3) of a reference multi-national cohort of children with healthy growth, while WLZ <-3 is characteristic of those with non-oedematous severe acute malnutrition (SAM)^2^. Stunting is the most prevalent form of childhood malnutrition, affecting around 150 million children with nearly one in three afflicted in parts of South Asia and sub-Saharan Africa^3^. Stunting often begins prenatally and is linked to maternal risk factors (height, age, education); after birth, it may continue to worsen under the influence of environmental stressors (e.g. poor diet, infection) until at least 2-years-of-age, whereafter LAZ scores remain low for children in many low-income countries^4–6^. Children under-5 who are stunted are five times more likely to die than non-stunted children and those who survive may suffer impairments in immune and cognitive development and have increased risk for chronic illnesses in adult life^7,8^.

Wasting and stunting are typically treated differently. Wasting is treated with high energy micronutrient-fortified ready-to-use supplementary or therapeutic foods (RUSF/RUTF) in the community or in hospital-based settings^9,10^. Approaches to treating stunting have been multifaceted, including strategies to reduce enteropathogen burden and environmental enteric dysfunction (EED)^11^ through access to clean water, sanitation, and improved hygiene (WASH), as well as various maternal, infant and young child feeding programs. To date evidence for the relative effectiveness of these approaches has been mixed^12,13^. Moreover, wasting and stunting frequently co-exist in the same infant/child, either concurrently or at different periods in their development, with co-occurrence being more common in severely wasted children and associated with significantly greater risk of mortality compared to either form of undernutrition alone^14–16^. Analyses of longitudinal studies of infant growth in the U.S., Honduras, Ghana and Malawi have found that weight gain in a preceding 3-month interval is associated with linear growth in the following interval^17,18^. A major challenge in interpreting these results is the lack of deeper knowledge of the underlying physiologic changes that occur within a given individual over time, and how these changes are related mechanistically to their growth trajectories.

Given the modest effectiveness of existing interventions, efforts are being made to obtain a more comprehensive view of the biological state or states associated with undernutrition, gather new insights into pathogenesis and identify additional therapeutic targets. One outcome of these efforts is recognition of the contributions of the gut microbiota. Birth cohort studies have disclosed that children with undernutrition have impaired postnatal development of their gut microbial communities compared to their age-matched healthy counterparts. This impaired development (microbiota immaturity) is worse in children with SAM compared to those with MAM and is not repaired by standard therapeutic food^19–21^. Phenotypic comparisons of gnotobiotic mice colonized with fecal microbiota from age-matched healthy children or those with wasting and stunting have revealed bacterial strains whose abundances/expressed functions are higher in healthy microbiota and that are able to ameliorate growth faltering when added to mice colonized with the microbiota from undernourished children^22^.

Based on these findings, gnotobiotic mice were used to ‘screen’ complementary foods commonly consumed in Bangladesh to test whether any had the ability to increase the fitness/expressed beneficial functions of growth associated taxa that are underrepresented during the weaning period in the microbiota of undernourished Bangladeshi children compared to their healthy counterparts. This effort ultimately led to the development of a ‘microbiota-directed complementary food’ prototype (‘MDCF-2’), containing ‘microbiota-active’ ingredients - chickpea, soy flour, peanut paste and green banana - along with vegetable oil, sugar and micronutrients^23^. A 3-month randomized controlled trial of MDCF-2 in 12-18-month-old Bangladeshi children with MAM revealed that children who received a MDCF-2 had a significantly greater rate of weight gain (WLZ) compared to children who received a standard RUSF, despite the fact that MDCF-2 had a 15% lower caloric density than the RUSF^24,25^. The superiority of MDCF-2 was linked to changes in the abundances of bacterial strains (metagenome-assembled genomes, MAGs) including strains of *Prevotella copri* that were significantly associated with improved WLZ and which possessed a distinct repertoire of polysaccharide utilization loci (PULs) compared to non-WLZ-associated *P. copri* MAGs^26^. The superior ponderal growth of children treated with MDCF-2 was accompanied by increases in the levels of plasma proteins that are biomarkers and mediators of musculoskeletal growth and neurodevelopment, plus decreases in the levels of plasma proteins that are biomarkers and mediators of inflammation^25^. Nonetheless, at the end of the 3-month intervention, children treated with MDCF-2 did not show statistically significant differences in their LAZ scores compared to those who received RUSF.

In this report, we describe the results of a 2-year follow-up of these children, which included anthropometry and assessment of the fecal microbiota and plasma proteome. Our results reveal that 3-months of MDCF-2 treatment not only provides a rapid and sustained improvement in ponderal growth, but also leads to superior linear growth in the follow-up period compared to RUSF. These findings, together with corresponding changes in the representation of growth associated MAGs and levels of plasma proteins involved in various facets of growth, provide evidence that the benefits of MDCF-2 treatment extend beyond the period of its administration in this population of Bangladeshi children with MAM.

## RESEARCH IN CONTEXT

### Evidence before this study

Undernutrition is the leading cause of death of children under 5-years-of-age worldwide. This multifaceted global health challenge is expected to become more severe in part due to intersecting effects of climate change, increasing economic disparities affecting low- and middle-income countries (LMICs), and continued geopolitical upheaval. Undernutrition not only increases the risk of death but is also associated with persistent sequelae including stunting, immune dysfunction, and cognitive deficits. Numerous epidemiologic studies have shown that undernutrition is not due to food insecurity alone. Moreover, current nutritional interventions, while reducing mortality, have had limited success in overcoming these sequelae.

### Added value of this study

We performed a 3-month randomized controlled trial in which a microbiota-directed complementary food formulation (MDCF) was compared to a standard RUSF in 12-18-month-old Bangladeshi children with MAM, who were also stunted. We previously reported that supplementation with MDCF improved ponderal growth compared to RUSF, despite the fact that the latter was more calorically dense. Here we report the results of a 2-year follow-up of these children which revealed significantly better LAZ scores in MDCF-2 compared to RUSF-treated children that was manifested only during the follow-up period. Improved LAZ was accompanied by significant differences in the representation of growth-associated bacterial taxa, plus significant elevations in plasma protein mediators/biomarkers of musculoskeletal growth and neurodevelopment that extended beyond the period of intervention.

### Implications of all the available evidence

Children with MAM and SAM have perturbed gut microbial community development that is manifested by alterations in the representation of growth-associated bacterial taxa as well as microbial genes comprising various metabolic pathways. The result of these perturbations are microbial communities whose configurations resemble those of chronologically younger children. Standard nutritional interventions in children do not fully repair these perturbed community states. Our findings suggest that microbiota-directed foods, designed to improve the representation and/or expressed metabolic functions of key gut bacteria, not only improve weight gain but also linear growth and plasma biomarkers of a range of developmental processes. These findings set the stage for additional studies to determine (i) the extent to which linear growth benefits of MDCF-2 are manifested in stunted children who are not also wasted; (ii) whether the observed plasma biomarker signatures/responses to MDCF-2 foreshadow clinical improvements in body composition and cognitive performance, and (iii) optimal conditions for initiating treatment (e.g., age, severity of undernutrition, MDCF dose/duration). Deeper knowledge of these relationships may enable an evolution in policies for treating undernourished children beyond current recommendations that are based largely on considerations of the energy density and macro-/micronutrient content of diets and therapeutic foods.

## METHODS

### Human study design

The study design for the randomized controlled trial comparing the efficacy of a microbiota-directed complementary food (MDCF-2) and a ready to use supplementary food (RUSF) in promoting weight gain in 12-18-month-old Bangladeshi children with MAM is shown in **Fig 1a**. The clinical study (ClinicalTrials.gov identifier:NCT04015999) was conducted in the Mirpur slum of Dhaka, Bangladesh from December, 2018 to October, 2019 as previously described^24,25^ The protocol was approved by the ethics review committee at the International Centre for Diarrhoeal Disease Research, Bangladesh (icddr,b). Children were screened and enrolled through household surveys in the community by Field Research Assistants and written consent was obtained from the parents of the study participants. Eligible children (male and female) with MAM (WLZ −2 to −3) who met the inclusion criteria were enrolled and randomly assigned to receive either a rice, lentil, milk powder, sugar and soybean oil-based RUSF, or MDCF-2, which contains chickpea flour, soy flour, peanut and green banana plus soybean oil and sugar. A total of 124 children were enrolled (n=62/arm)^24^. At enrollment (baseline), anthropometric assessments were performed, and fecal and plasma samples were collected from each child. MDCF-2 and RUSF, freshly prepared at the icddr,b Food Processing Facility located in Mirpur were provided to participants twice daily for 3 months under supervision by trained study staff (morning and afternoon, 2 x 25g servings/day). The total energy provided by the supplements was 220-250kcal/day or ∼20-25% of the daily recommended energy intake (and 70% of the recommended micronutrients) for healthy 12–18-month-old children. Caregivers were instructed not to feed their children during the 2-hour period before each visit; otherwise, their usual breastfeeding and complementary feeding practices were maintained. Fecal specimens were collected prior to intervention, weekly during the 1st month of intervention, and every 4 weeks during the second and third month of intervention. After cessation of the intervention, fecal sampling was performed at the 1, 6 and 12 months of follow-up. (Note that these follow-up time points relate to the end of *study months* 4, 9 and 15 respectively; **Fig. 1a**). Blood samples were collected prior to intervention, at the end of first and third month of intervention, and at 1, 6 and 12 months of follow-up.

**Figure 1.**
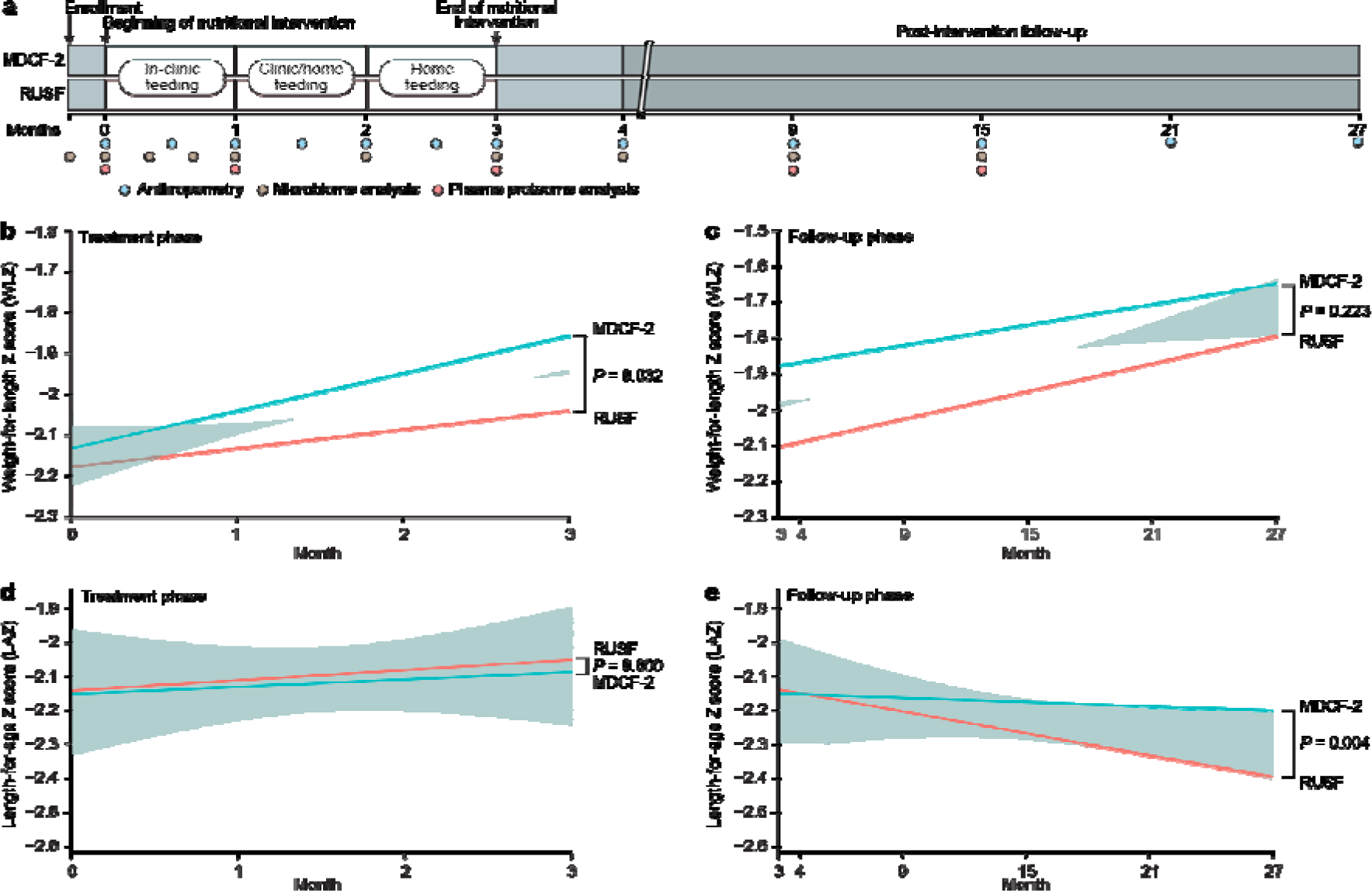
Anthropometric trajectories during treatment with MDCF-2 or RUSF and during the two year post-intervention follow-up period. (**a**) Trial design, including timing of biospecimen collection for analysis of the microbiome and plasma proteome. (**b,c**) Weight-for-length Z-score (WLZ) trajectories during the three months of MDCF-2 or RUSF treatment (panel b) and during the two years of post-intervention follow-up (panel c). (**d,e**) Length-for-age Z-score (LAZ) trajectories during the three months of MDCF-2 versus RUSF treatment phase of the trial (panel d) or the two years of post-intervention follow-up (panel e). Each plot indicates a simple linear fit of the data for the MDCF-2 (blue) and RUSF (red) treatment groups along with the 95% confidence interval. The linear, mixed effects model for each anthropometric assessment (Eq. 2; *Methods*) was used to determine the coefficient (β) and statistical significance (*P*) for the specified effect. For the treatment phase analysis, the beginning of nutritional intervention was used at the baseline measurement, whereas for the follow-up analysis, the baseline used was the end of 3-month nutritional intervention.

Anthropometric assessments were performed by trained field research assistants at enrollment (just prior to treatment), every 15 days during the intervention period, 1 month after cessation of treatment and at 9, 15, 21 and 27 months after enrollment into the study. The presence/absence of any illness during the 7 days preceding each anthropometric assessment was recorded. The weight of each child was determined using a digital scale with 2 g precision (Seca, model 728, Germany). Length was measured using an infantometer (Seca, model 416, Germany), and mid-upper arm circumference (MUAC) was determined to the nearest millimeter (using a non-stretch tape). Each measurement was taken twice at a single timepoint by two separate individuals and the average score was recorded. Z scores were calculated using the new child growth standards of the World Health Organization (WHO)^2^. For each of the four main anthropometric assessments, we modeled the treatment group-specific trajectories using the following linear mixed effects model:

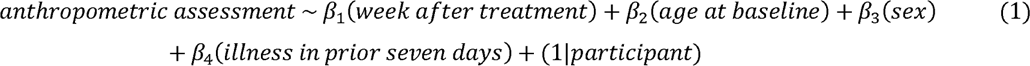

We employed the model below to compare effects between the treatment groups:

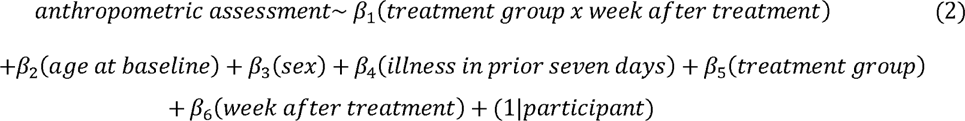

### Biospecimen collection

Fecal and plasma specimens were collected as previously described^25^. In brief, within 20 minutes of defecation, aliquots of the fecal sample were transferred by the field worker into sterile 2LmL cryovials and transferred into cryo-shippers (Taylor Wharton/ Worthington Industries CX300) that had been pre-charged with liquid nitrogen. Cryo-shippers were transported to the study center where their contents were recorded, and the vials transferred to a −80°C freezer. Venous blood was collected in EDTA-plasma tubes; plasma recovered after centrifugation (3000 x g for 10 minutes at room temperature) was aliquoted into cryovials and stored at −80°C. Specimens were shipped to Washington University in St Louis USA on dry ice and stored at −80°C in a dedicated biospecimen repository prior to analysis, with approval from the Washington University Human Research Protection Office. For this study, 225 fecal samples (n=114 at 6 months follow-up and n=111 at 12 months) and 219 plasma samples (n=111 at 6 months follow-up and n=108 at 12 months) were collected/analyzed. Additional fecal and plasma samples included in the analyses described here were collected prior to, during and at the end of the intervention, plus at the 1-month follow-up time point, as previously reported^25,28^.

## Quantification of enteropathogens by multiplex qPCR

The same qPCR assay employed in the treatment phase of the study was applied in the post-treatment follow-up phase to measure 23 bacterial, viral and protozoan enteropathogens^25–27^. Enteropathogen abundances were compared between the treatment groups at the end of treatment, and at 6- and 12-months post-treatment using both Kruskal-Wallis tests applied to log_10_-transformed enteropathogen abundances at each sampling timepoint and linear, mixed-effects models of the following form:

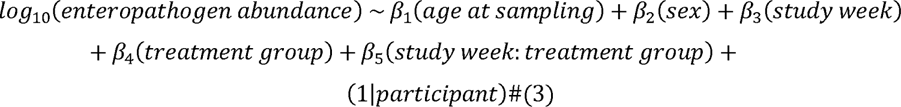

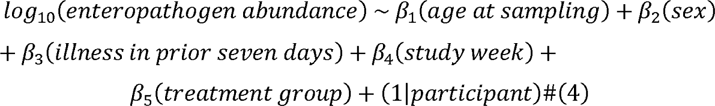

Significance for the Kruskal-Wallis test, Eq. 3 (β_5_ term) and Eq. 4 (β_3_ term) was defined by q<0.05 (*P-value* adjusted to control for multiple hypotheses using the Benjamini-Hochberg correction^29^).

### Isolation and short-read shotgun sequencing of fecal DNA

DNA was isolated from 225 fecal samples representing the 6- and 12-month post-treatment follow-up timepoints as previously described^28^ and shotgun metagenomic sequencing libraries were prepared with a modified Nextera XT (Illumina) protocol^30^. Libraries were quantified, balanced, pooled, and sequenced to a depth of 8.96×10^7^±2.09×10^7^ 150 nt paired-end reads/sample (mean±SD) at similar scale and on the same Illumina NovaSeq 6000 sequencing platform (S4 flow cell) as the treatment phase fecal metagenomic sequencing effort described previously^28^. [Negative (water only) process controls were included in both the treatment phase and follow-up sequencing efforts. Library preparation and sequencing of these samples yielded 6.70×10^4^±3.01×10^4^ (mean±SD) reads]. Reads were demultiplexed and pre-processed to remove low-quality bases/reads, as well as reads mapping to the hg19 assembly of the human genome^28^.

### Quantitation of Abundances of Metagenome Assembled Genomes (MAGs)

Pre-processed shotgun data from the 6- and 12-month follow-up time points were mapped using Kallisto (v0.43.0)^31^ to a single quantification index from the set of 1,000 high-quality MAGs (completeness ≥90%, contamination <5%) generated from data collected during the treatment phase and 1 month follow-up time point of this study^28^. The 1,000 MAGs were produced using an assembly strategy in which shotgun sequencing reads generated from fecal samples serially collected from a given trial participant were aggregated and assembled into a ‘participant-specific’ MAG set. MAGs from all participants were then compared and dereplicated to generate a set of study-wide MAGs.

MAGs were considered redundant if they shared 99% average nucleotide identity (ANI)^28^. Taxonomy was subsequently assigned to these MAGs using a consensus approach based on (i) Kraken (v2.0.8)^32^ and Bracken (v2.5)^33^ plus (ii) the Genome Taxonomy Database Toolkit (GTDB-tk)^34^ and GTDB database (v0.95). We then mapped fecal shotgun sequencing data from the additional follow-up time points to the 1,000 MAGs assembled previously to facilitate the merging of the datasets. After merging, the dataset contained abundance data for each of the 1,000 MAGs across 928 samples (703 samples representing the period from pretreatment through 1 month of follow-up^28^ and 225 from the 6- and 12-month follow-up time points) with matching anthropometric assessments. This dataset was then filtered to exclude MAGs below a threshold count of 5 reads per kilobase per million in ≥ 40 % of samples, yielding a final dataset containing 851 MAGs.

### Principal components analysis

We assayed the stability of the microbiota over the first 12 months of post-intervention follow-up using principal components analysis (PCA) applied to the filtered, VST-transformed dataset spanning treatment and follow-up samples described above. PCA was performed using the ‘prcomp’ function in R (v4.3.2); sample projections and information regarding variance explained by each principal component (PC) were extracted as previously described^28^. PCs explaining variance above ‘background’ were correlated to the age at which each fecal sample was collected (Pearson) (background defined by applying PCA to a permuted dataset^28^). PC projections were compared over time and between treatment groups using t-tests, with statistical significance defined as *P-*value < 0.05.

### Identifying MAGs and MAG features associated with MDCF-2 treatment and/or anthropometric recovery

We used linear mixed effects modeling (dream, part of the variancePartition package v1.24.1, in R v4.3.2)^35^ to relate the untransformed abundances of MAGs to ‘treatment group’, ‘weeks after intervention’ and the interaction of these terms, while controlling for repeated sampling from each participant:

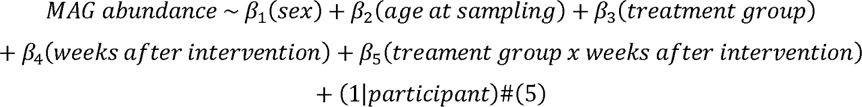

This model was applied to the abundance dataset containing 851 MAGs in fecal samples collected at the end of treatment plus 1, 6, and 12 months into the post-intervention follow-up (defined in the model as 12, 16, 36 and 60 weeks from the start of the trial). Relationships between MAG abundances and terms in the model were considered significant if they displayed q<0.05.

We employed gene set enrichment analyses (GSEA; fgsea v3.14)^36^ to identify similar abundance responses in the set of 75 positively or 147 negatively WLZ-associated MAGs identified in our prior analyses of treatment-phase fecal samples^28^. MAGs were ranked based on a standardized z statistic describing their relationship with terms from the linear mixed effects model described in Eq. 5 above, after which GSEA was performed. Sets of MAGs displaying q<0.05 were considered significant.

We also performed a differential abundance analysis of MAGs between the MDCF-2 and RUSF treatment groups at each time point using a simplified model (dream, v1.24.1) relating MAG abundance only to the treatment group, with significant differences between groups defined as having a q<0.05. Enrichment analyses for the positively or negatively WLZ-associated MAGs were also applied to comparisons of MAG abundances at each time point using the methods and statistical criteria described above.

To identify MAGs whose abundances were associated with the rate of change in LAZ in the follow-up phase, we transformed the filtered MAG abundance dataset using the Variance Stabilizing Transformation (VST)^37^; we then filtered this dataset to the timeframe spanning the end of treatment and 12-months of follow-up and determined relationships with LAZ using the following model:

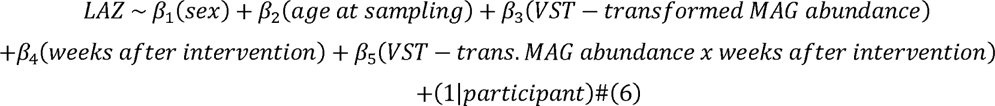

Statistical significance was determined for the β_3_ term using ANOVA with a cutoff of q<0.05. We additionally used the β_3_ coefficients corresponding to each MAG abundance relationship with LAZ as a ranking factor for GSEA, after which sets of MAGs, grouped by (i) species-level taxonomic assignment, (ii) WLZ-association in the treatment phase and (iii) the presence or absence of encoded metabolic pathways were tested for enrichment (significant enrichments were indicated by q<0.05). Metabolic pathway annotations for MAGs were based on alignment to the mcSEED database and *in silico* prediction of pathway presence/absence as previously described^28^.

### Aptamer-based plasma proteomic analysis

The SomaScan proteomic assay plasma/serum kit (SomaLogic, v4.0) was used as previously described^25^ to quantify 5,284 proteins in the plasma samples that had been collected from study participants at enrollment and 1, 3, 9 and 15 months after enrollment. Plasma samples were separated into two batches for data acquisition, with the first consisting of samples collected at enrollment, 1- and 3-months (treatment phase), and the latter from 9- and 15-months (follow-up). Each 50 µL aliquot of plasma was incubated with NHS-biotin-tagged aptamer probes (‘SOMAmers’) to form protein-specific SOMAmer complexes. After immobilization on streptavidin beads, complexes were cleaved, denatured, eluted, and hybridized to customized Agilent DNA microarrays. Arrays were scanned (Agilent SureScan; 5 µm resolution) and the Cy3 fluorescence signal processed using SomaLogic’s standardization procedures^25^. Protein abundances were log_2_-transformed prior to analysis.

To assess whether the 75 plasma proteins that were associated with the rate of weight gain (β-WLZ) in the treatment phase^25^ were also predictive of the rate of change in WLZ or LAZ during the follow-up period, we first calculated the rate of change of these 75 proteins during treatment (0-3 months) for each participant in each treatment arm [n=113 samples at enrollment, n=114 at the end of treatment]. Next, we calculated the rate of change in WLZ and LAZ in each participant during the follow-up period (months 3-27) and used linear models to relate the rate of change of each protein during treatment to the rate of change of LAZ or WLZ during the follow-up period:

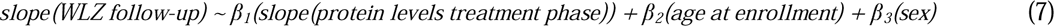

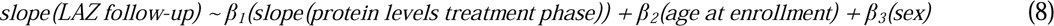

Significance was assigned after correcting for multiple testing for each anthropometric measure (q<0.1).

We applied a linear mixed effects model to test whether any of the proteins significantly associated with anthropometric changes (as defined from Eq. 7 and 8) exhibited a greater rate of change in one or other treatment group during the treatment phase:

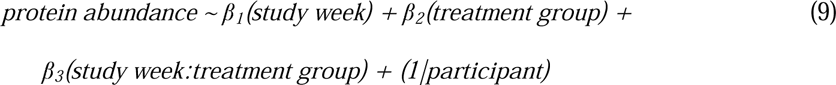

ANOVA was used to test for statistical significance, with correction for multiple testing (q<0.1). To determine if the resulting proteins were significantly associated with a treatment group as a whole, we applied a one-way t-test to the β_3_(study week:treatment group) coefficients collected for each protein.

For plasma samples collected at the 6- and 12-month follow-up time points [n=111 samples at month 6 and n=108 at month 12], a linear model was applied to identify LAZ-associated plasma proteins whose levels were differentially abundant between the two treatment arms at each time point:

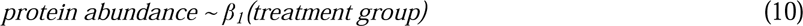

We applied a one-way t-test to the β_1_(treatment group) coefficients for proteins at the 6- and 12-month follow-up time points and subsequently applied a false-discovery rate correction on the *P*-values obtained from the t-tests applied to Eq. 9 *β_3_* and Eq. 10 *β_1_* coefficients (q<0.05).

### Illness and dietary intake during the follow-up period

Each child was monitored for illness daily for one continuous week ending at or near the time of anthropometry data collection at the end of treatment and at the end of 1, 6, 12, 18, and 24 months of post-intervention follow-up. Food frequency questionnaires were administered at the same time points and used to calculate Minimum Acceptable Diet (MAD) per World Health Organization and UNICEF guidelines^38^. MAD is only valid for children 6-23 months old. Children in our clinical trial surpassed this range between 1- and 6-months of post-intervention follow-up. Therefore, we applied chi-squared tests comparing whether MAD criteria were met for the MDCF-2 versus the RUSF treatment groups at the 1- and 6-month follow-up timepoints. Significance was defined by *P*<0.05.

## RESULTS

### Anthropometric recovery of children with MAM during and after treatment with MDCF-2

Out of 124 children, 118 (n=59/arm) completed the intervention; their anthropometric characteristics at the end of the treatment phase (i.e., the beginning of follow-up) are shown in **Table 1**. For each time point of anthropometric assessment (**Figure 1a**), we modeled the treatment group-specific trajectories as well as differences between groups using linear mixed effects models, controlling for, treatment time, age, sex and the presence of any illnesses in the 7 days prior to anthropometry (see *Methods,* Eq. 1 and 2; **Supplementary Table S1A,B**). The superior WLZ achieved by children in the MDCF-2 arm at the end of treatment (**Figure 1b**) was maintained through follow-up, though the difference in rate of change of WLZ between the groups was no longer significant (*P*=0.22; linear mixed effects model) (**Figure 1c; Supplementary Table S1B**). Weight-for-age (WAZ) also trended higher in the follow-up period for MDCF-2 treated children, although this difference was not statistically significant (**Supplementary Table S1B**).

**Table 1:**
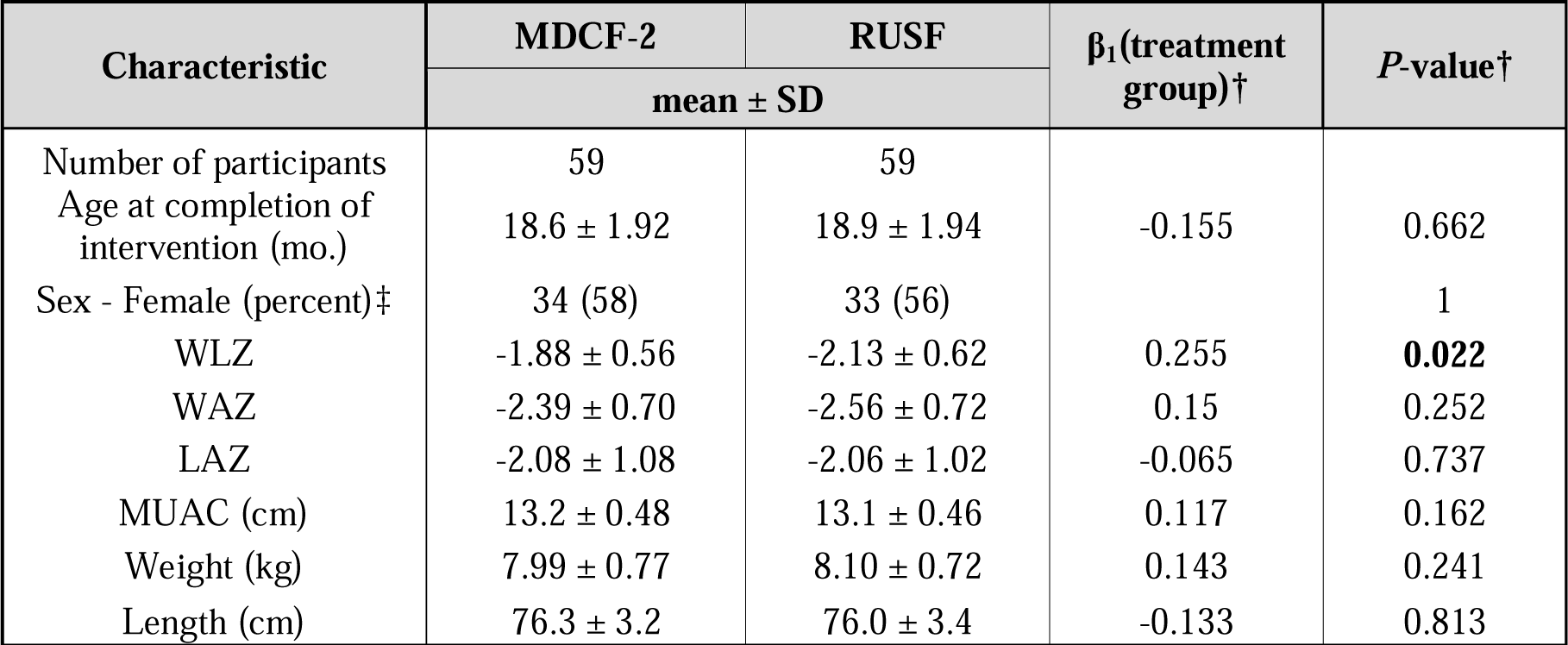
Anthropometric characteristics of children in each study arm at the end of the 3-month intervention (beginning of follow-up phase) † Model: anthropometry ∼ β_1_(treatment group) + β_2_(age at baseline) + β_3_(sex) + β_4_(illness in prior 7 days). Note that the ‘age at baseline’ term was omitted when ‘age at baseline’ was tested. ‡ Fisher’s Exact Test.

| Characteristic | MDCF-2 | RUSF | β<sub>1</sub>(treatment group)† | P-value‡ |
| --- | --- | --- | --- | --- |
|  | mean ± SD |  |  |  |
| Number of participants | 59 | 59 |  |  |
| Age at completion of intervention (mo.) | 18.6 ± 1.92 | 18.9 ± 1.94 | -0.155 | 0.662 |
| Sex - Female (percent)‡ | 34 (58) | 33 (56) |  | 1 |
| WLZ | -1.88 ± 0.56 | -2.13 ± 0.62 | 0.255 | **0.022** |
| WAZ | -2.39 ± 0.70 | -2.56 ± 0.72 | 0.15 | 0.252 |
| LAZ | -2.08 ± 1.08 | -2.06 ± 1.02 | -0.065 | 0.737 |
| MUAC (cm) | 13.2 ± 0.48 | 13.1 ± 0.46 | 0.117 | 0.162 |
| Weight (kg) | 7.99 ± 0.77 | 8.10 ± 0.72 | 0.143 | 0.241 |
| Length (cm) | 76.3 ± 3.2 | 76.0 ± 3.4 | -0.133 | 0.813 |

Linear growth faltering often begins *in utero* and may continue to worsen during the first 2 postnatal years in low-income countries such as Bangladesh^4^. In our randomized controlled trial, LAZ was not different between the two groups during the treatment phase (*P*=0.6 linear mixed effects model; **Figure 1d, Supplementary Table S1A**). However, in the 24-month post-intervention follow-up period (**Table 2**), we observed a statistically significant decrease in the rate of LAZ decline in children that had received 3 months of MDCF-2 supplementation compared to those that had received RUSF (*P*=0.004; **Figure 1e; Supplementary Table S1B**). To examine whether differences in the quality of diets consumed by children in the follow-up period may have contributed to differences in linear growth trajectories between the intervention groups, we examined food frequency data collected during follow-up period and compared the Minimum Acceptable Diet (MAD) scores for each group. MAD is an indicator of infant and young child feeding (IYCF) practices for 6-23-month-old children that is used to describe the quality of a child’s diet and is defined as the percentage of children (in the 6–23-month age range) who consumed a minimum acceptable diet during the previous day^38^. MAD was not significantly different between the MDCF-2 and RUSF groups at the 1- and 6-month follow-up time points when this measurement was applicable based on age (chi-squared *P*>0.05, **Supplementary Table S1C**). Taken together, these results indicate that compared to the more calorically dense RUSF used in this RCT of children with MAM, a relatively short intervention with MDCF-2 provided improvements not only in their ponderal growth, but also a sustained difference in linear growth that is not explained by differences in quality of diet consumed by children during follow-up.

**Table 2:** LAZ scores of participants in each study arm during the 24-month follow-up period.

| Months of follow-up<br>after intervention | MDCF-2 (n=59) | RUSF (n=59) | Difference (95% CI) |
| --- | --- | --- | --- |
|  | LAZ (mean ± SD) |  |  |
| 1 | -2.09 ± 1.07 | -2.09 ± 1.00 | - |
| 6 | -2.28 ± 1.00 | -2.31 ± 0.92 | 0.03 (-0.32, 0.39) |
| 12 | -2.24 ± 0.93 | -2.42 ± 0.86 | 0.18 (-0.16, 0.52) |
| 18 | -2.26 ± 0.96 | -2.42 ± 0.87 | 0.16 (-0.18, 0.51) |
| 24 | -2.09 ± 0.92 | -2.22 ± 0.80 | 0.13 (-0.20, 0.46) |

### Effects of MDCF-2 on the microbiota

We next assessed whether the improvements in anthropometry during the follow-up period were associated with durable changes in the microbiota resulting from treatment with MDCF-2. Of the 118 children who completed the intervention, we collected fecal samples from 114 after 6 months of follow up (n=57/treatment group), and 111 at 12 months (n=56, MDCF-2 treatment group; n=55, RUSF treatment group) (i.e., there was only 6% loss to follow-up at 12 months).

We employed a qPCR assay^25^ to quantify the abundances of 23 bacterial, viral and protozoan enteropathogens in each child’s fecal samples (**Supplementary Table S2A**). Neither pairwise comparisons of enteropathogen abundances, nor the total number of detected enteropathogens at the cessation of treatment, or at 6- or 12-months post-treatment revealed any significant differences between the treatment groups (Kruskal-Wallis tests, q>0.05). Likewise, linear mixed-effects models designed to identify the effects of interactions between treatment group and time after cessation of treatment (controlling for age and sex) on enteropathogen abundances failed to identify significant differences between the treatment groups (**Supplementary Table S2B**). We subsequently examined the relationship between the enteropathogen levels and illnesses reported in the seven days prior to fecal sample collection; these analyses revealed a significant positive relationship between diarrheal illness and (i) the total number of enteropathogens detected and (ii) the abundances of *C. jejuni*, enterotoxigenic *E. coli*, Cryptosporidium and diarrheal illness, independent of age, sex, timepoint or treatment group (**Supplementary Table S2C**).

To analyze the fecal microbial communities of MDCF-2- and RUSF-treated participants sampled during the post-intervention follow-up period, we mapped shotgun sequencing data generated from fecal samples collected at the 6- and 12-month follow-up timepoints to the set of 1,000 high-quality metagenome assembled genomes (MAGs) assembled from fecal shotgun sequencing data collected during the treatment phase of the study^28^. These datasets were merged with data generated from the treatment phase of the study and one-month follow-up, after which filtering was applied to exclude sparsely represented MAGs. This effort resulted in a dataset comprising 851 MAGs across 928 samples with matched anthropometry data (see *Methods*).

We applied a principal components analysis (PCA) to the full MAG dataset spanning pretreatment through 12 months of follow-up. We identified principal components (PCs) that explained variance above background (*Methods*). PC1, explaining 19.2% of variance, was significantly correlated with a child’s age at the time of sample collection (Pearson *P*<2.2×10^−16^). We therefore used the projection of each sample along PC1 as a dimensionality-reduced metric of microbiota ‘age’. As expected, given that the treatment and early follow-up phases of the study occurred in children at an age when the microbiota is developing^19^, there was a significant change in position along PC1 between samples collected at 1 and 6 months and 6 and 12 months of follow-up (*P*=1.87×10^−9^ and *P*= 6.43×10^−7^, respectively; t-test). However, we observed no significant difference between the PC1 projections of samples obtained from participants in the MDCF-2 and RUSF arms during the follow-up phase, when children were between 16- and 33-months-old (*P*>0.05, t-tests).

We proceeded to analyze the abundances of each individual MAG over time during the period between the end of treatment and 12 months of follow-up in children formerly treated with MDCF-2 or RUSF. As in the treatment phase, no single MAG met our criteria for statistically significant differential abundance or differential rate of change of abundance between the two treatment groups during this period (q>0.05, linear mixed effects model, Eq. 5; **Supplementary Table S3A**). However, employing gene set enrichment analysis (GSEA), we compared the representation of the MAGs previously identified^28^ as significantly positively (n=75) or negatively (n=147) associated with WLZ during the treatment phase, in fecal samples obtained at the end of treatment and at 1, 6 and 12 months of follow-up. MAGs that were positively associated with WLZ were enriched in the MDCF-2 treatment group, while MAGs that were negatively associated with WLZ were enriched in the RUSF group at the end of treatment, and at one month of follow-up (GSEA q<0.05, (**Fig. 2a**, **Supplementary Table S3C,D**). The enrichment of positively WLZ-associated MAGs was driven by 15 MAGs assigned to the species *Agathobacter rectalis* (5 MAGs), *Blautia massiliensis* (5 MAGs), *CAG-217 sp00436335* (family Acutalibacteraceae, 2 MAGs), *ER4 sp000765235* (family Oscillospiraceae, 2 MAGs), *Parolsenella catena* (2 MAGs), and *Prevotella copri* (2 MAGs; **Fig. 2b**). Notably, these two *P. copri* MAGs were Bg0018 and Bg0019 – both positively associated with WLZ - that express polysaccharide utilization loci involved in the metabolism of glycans enriched in MDCF-2 compared to RUSF^28^. The enrichment of this set of 15 MAGS was no longer statistically significant at the 6-month post-intervention time point (GSEA q>0.05).

**Figure 2.**
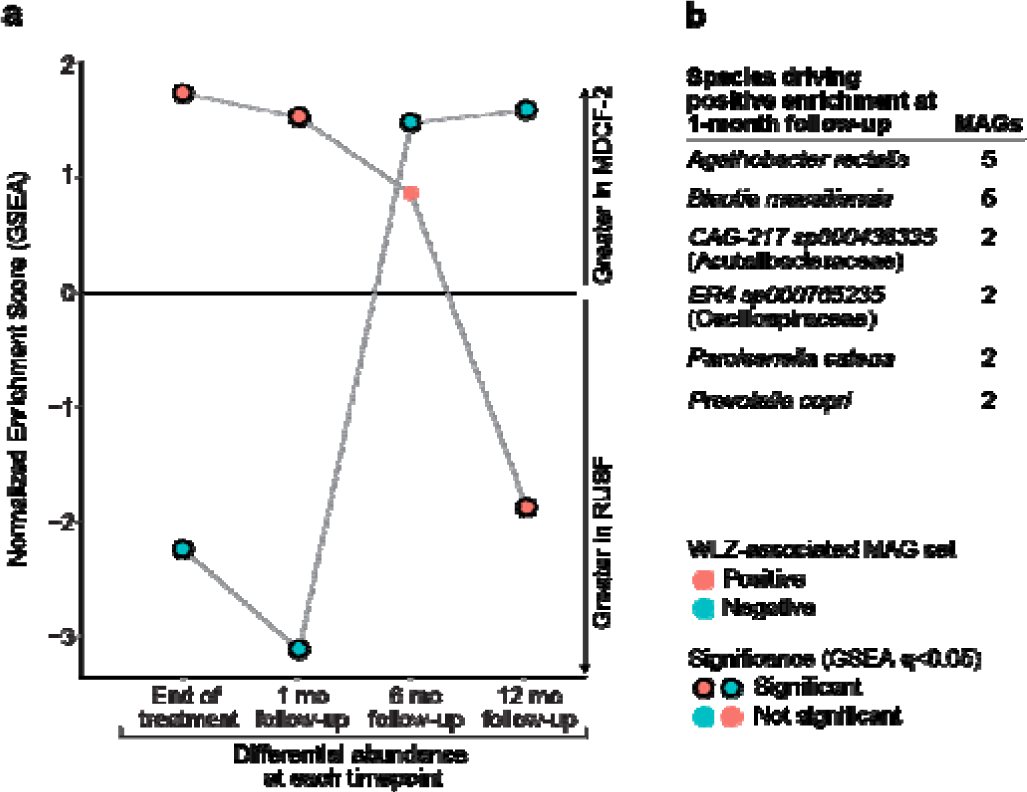
Differences in bacterial MAG abundances between fecal samples collected from MDCF-2- and RUSF-treated trial participants during the post-intervention follow-up. (a) The enrichment of groups of positively (red) or negatively (blue) WLZ-associated MAGs after ranking by differential abundance between the MDCF-2 or RUSF groups at end of treatment and at the 1-, 6-, or 12-month post-intervention time points. Normalized Enrichment Scores (NES) were determined by GSEA. Points representing significant NES significant (q<0.05) are outlined in black. (**b**) Taxonomic assignments of MAGs driving the enrichment of positively WLZ-associated MAGs in the analysis depicted in (a). Species assignments are shown for taxa containing >1 MAG.

Given the effect of prior MDCF-2 treatment on LAZ in the follow-up period, we initially used the same approach we had employed to identify WLZ-associated MAGs to search for LAZ-associated MAGs in the follow-up phase. Our efforts to model the relationship between MAG abundance and LAZ yielded only one significantly associated MAG, Bg0899, which was negatively associated and classified within the family Lachnospiraceae (q<0.05, linear mixed effects model (Eq.4), **Supplementary Table S3E**). Therefore, we expanded this analysis by ranking MAGs based on the coefficient describing their relationship with LAZ over time in the follow-up period [β_3_(MAG abundance x weeks after intervention)] and applying GSEA to sets of MAGs defined by their species assignments. Using this approach, we identified a group of four species (*Agathobacter faecis*, *Blautia massiliensis*, *Lachnospira sp000436475* and *Dialister sp000434475* and) that were positively associated with LAZ (q<0.05, GSEA, **Supplementary Table S3F**). *Agathobacter* produce the beneficial short chain fatty acid (SCFA) butyrate though fermentation of mono-di- and oligo-saccharides^39^, while *Blautia massiliensis* is reported to be enriched in carbohydrate transport and metabolism capacities compared with other species in the genus *Blautia*, with acetate a key product of fermentation^40,41^. *Lachnospira* and *Dialister* also contribute to the fermentation of carbohydrates and production of SCFA.

Finally, we looked for relationships between MAGs that were significantly associated with WLZ in the treatment and follow-up phases of the trial. This analysis revealed a group of 75 MAGs positively correlated with WLZ in the treatment phase^28^ that were also enriched among LAZ correlated MAGs during follow-up (q=1.24×10^−4^, GSEA applied to MAGs ranked by their LAZ association; **Supplementary Table S3G**). Notably, the 25 MAGs that had the strongest associations with both LAZ and WLZ were predominantly assigned to the families Lachnospiraceae (n=12 MAGs), Ruminococcaceae (n=8) or Oscillospiraceae (n=3) (**Supplementary Table S3H**). An analysis of MAGs whose abundances were positively associated with LAZ yielded no significantly enriched metabolic pathways (GSEA, q>0.05). However, DNA abundance is an imprecise proxy for functional activity and additional analyses involving microbial RNA-Seq are required to characterize the relationship between *expressed* community functions influenced by MDCF-2 and linear growth.

### Effects of MDCF-2 on the plasma proteome

We next sought to determine whether the plasma proteins positively (n=70) or negatively (n=5) associated with improvements in weight gain (β-WLZ) during the treatment phase [n=113 plasma samples analyzed at enrollment and n=114 at the end of treatment]^25^ were also associated with the rate of change in either WLZ or LAZ during follow-up. We used linear models, accounting for age and sex (Eq. 7,8) to regress the rate of change of each of the 75 proteins to the rate of change of WLZ or LAZ during the 24-month follow-up phase. This analysis disclosed that while none of the proteins remained significantly associated with ponderal growth (WLZ) (**Supplementary Table S4A**), 37 were significantly positively associated with rate of change of LAZ during follow-up (q< 0.1, **Fig. 3a Supplementary Table S4B**); all of these were positively associated with WLZ during the treatment phase^25^. They include biomarkers/mediators of musculoskeletal development; e.g., cell adhesion molecule-related/down-regulated by oncogenes (CDON) which is a positive regulator of myogenesis^42^, dermatopontin (DPT) which accelerates collagen fibrillogenesis^43^, thrombospondins-3 and −4 (THBS3,4) which are involved in tissue remodeling and skeletal maturation via interaction with the extracellular matrix^44,45^, plus a number of collagens (e.g., COL15A1, COL9A1, COL6A3). Other notable proteins among the 37 positively associated with LAZ include insulin-like growth factor-1 (IGF-1) which acts in concert with growth hormone to control linear growth in children^46^ and a Wnt inhibitor, secreted frizzled-related protein 4 (SFRP4), involved in formation of bone cortex^47^. The list also includes effectors of CNS development: e.g., RET which encodes the receptor tyrosine kinase for glial cell derived neurotrophic factor (GDNF)^48^; the axon guidance protein SLIT and NTRK like family member 5 (SLITRK5)^49^, BDNF/NT-3 growth factors receptor (NTRK2) which is involved in synaptogenesis and learning and memory^50^, plus roundabout homolog 2 (ROBO2), an axon guidance receptor recently reported to also promote osteoblast differentiation and mineralization^51^ (see **Fig. 3b** for complete list of these proteins and their functional annotations).

**Fig 3.**
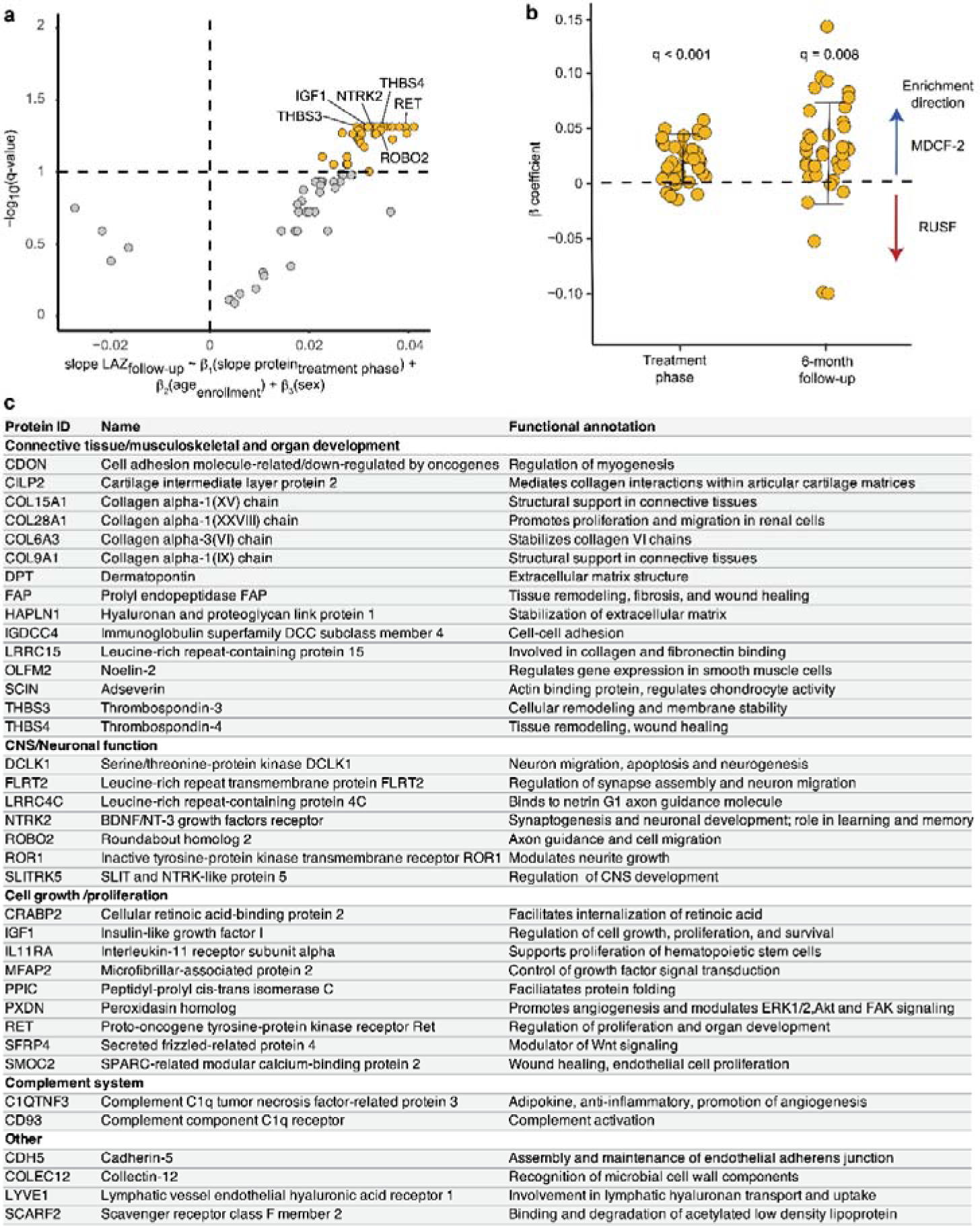
- Effects of MDCF-2 on the plasma proteome of children with MAM during nutritional intervention and in the follow-up period. (a) Volcano plot of WLZ-associated plasma proteins that are also associated with LAZ responses during the post-treatment follow-up period. The linear model described under the plot was applied to relate the rate of change for each participant in the levels of proteins during the treatment-phase to the rate of change of LAZ during the follow-up. Significant LAZ-predictive proteins (q<0.1) are colored in orange. (**b**) Annotated functions of the 37 plasma proteins associated with the effect of MDCF-2 on LAZ during the follow-up period. (**c**) Association of the 37 proteins with treatment arm during the 3-month treatment phase and at the 6-month follow-up time point. One-way t-tests were applied to the β3 coefficient in Eq. 9 and the β1 coefficient in Eq. 10 (see *Methods*, q<0.05) at each time point to test the statistical significance of their enrichment in either the MDCF-2 or RUSF treatment arm.

For each of the 37 LAZ-associated proteins, we applied a linear mixed-effects model (Eq. 9 in *Methods*) to obtain their differential rates of change in levels during the treatment phase between the MDCF-2 and RUSF groups (**Supplementary Table S4C**). A one-way t-test was then performed on the β*_3_(month:treatment group)* coefficients; each coefficient represents the differential rate of change in the level of each protein between the two groups during treatment. The analysis revealed that these proteins, as a group, were significantly enriched in the MDCF-2 arm (FDR corrected q<0.05; **Fig 3c**). We next used a linear model (Eq. 10 in *Methods*) to test whether the LAZ-associated proteins were differentially abundant between the MDCF-2 and RUSF treatment arms at month 6 and month 12 of the follow-up period [n=111 plasma samples analyzed at the end of month 6; n=108 at the end of month 12] (**Supplementary Table S4D,E**). While none of the 37 proteins exhibited significant differences in their levels between treatment arms at either follow-up time point (q>0.1), as a group their levels in plasma was significantly higher in MDCF-2 compared to RUSF treated children at the 6-month follow-up time point [(one-way t-test applied to the β*_1_(treatment group)* coefficients in Eq. 10 with q<0.05 (**Fig. 3c**)]. By 12 months of follow-up, this enrichment was no longer evident. Taken together, these results indicate that the 3-month intervention with MDCF-2 in 12-18-month-old Bangladeshi children with MAM improved ponderal and linear growth and produced increases in the levels of a group of plasma biomarkers/ mediators of growth and CNS development that were evident for at least 6 months after cessation of treatment.

## Discussion

In this study, we performed a two-year follow-up of a cohort of 12-18-month-old Bangladeshi children who had been treated for moderate acute malnutrition (MAM) for 3 months with either a microbiota-directed complementary food (MDCF-2), or a standard ready-to-use supplementary food (RUSF)^25^. We assessed their anthropometric recovery and quantified changes in the representation of >800 bacterial strains (MAGs) present in their fecal microbial communities, plus levels of plasma proteins during the follow-up period. WLZ scores, which were significantly improved by MDCF-2 compared to RUSF at the end of treatment, remained higher in the MDCF-2 group during follow-up. Moreover, despite there being no significant differences in LAZ between the groups at the end of the 3-month intervention, there was a significant difference between the groups during follow-up; namely, the rate of decline in LAZ, which is commonly seen in children in the first few years of life in resource-poor countries in South Asia and Africa^4,5^, was significantly less in children who had received MDCF-2 in the antecedent treatment phase. There were no significant differences in diet quality or enteropathogen burden between the treatment groups during the follow-up period. We identified a number of MAGs that were correlated with LAZ after cessation of the intervention, including strains capable of fermenting dietary glycans to short chain fatty acids. Moreover, several MAGs belonging to Lachnospiraceae, Ruminococcaceae and Oscillospiraceae were linked to both WLZ during treatment and LAZ during follow-up.

The improved linear growth in MDCF-2-treated children in the follow-up period was associated with elevated levels of a group of 37 plasma proteins that persisted for at least 6 months after cessation of treatment; these proteins were previously identified as being positively correlated with WLZ during the treatment phase of the study^25^; their functions are linked to various facets of healthy growth, including musculoskeletal development, central nervous system development and cell growth/proliferation. This set of plasma proteins represent candidate ‘leading indicators’ of linear growth recovery in undernourished Bangladeshi children. Moreover, they provide a rationale for including more specific clinical measurements beyond anthropometry to more comprehensively characterize physiologic responses to MDCF-2; for example, musculoskeletal and cognitive development which may be manifest over longer time frames than changes in WLZ (or LAZ).

There is a paucity of sufficiently powered longitudinal studies and randomized controlled trials that incorporate detailed characterization of the functional capacity of the microbiome, such as shotgun sequencing of fecal DNA to define changes in the representation of MAGs, and microbial RNA-Seq to relate the expression of metabolic pathways and their products to clinical outcomes. One recent example of the importance of this kind of approach employed a metagenomic analysis of fecal samples from 335 infants/young children living in rural Zimbabwe where there is a high prevalence of stunting. This study revealed that genomic features of the fecal microbial community, in particular, the representation carbohydrate degradation pathways (rather than taxonomic representation), were predictive of linear growth of children during the period of complementary feeding^52^. This finding is consistent with our own studies of growth responses to MDCF-2 in children with MAM; namely that improvements in ponderal growth are driven by specific strains (MAGs) of *P. copri* that express the appropriate repertoire of carbohydrate active enzymes (CAZymes) required to utilize MDCF-2 glycans^28^. Additional studies incorporating similar types of analyses (microbial gene expression, fecal metabolomics) will be required to characterize the *functional* maturation of the microbiota, as well as the relationship between changes in functional activity that link responses to MDCF-2 with linear growth outcomes (including the durability of changes in metabolic activities both during and following intervention).

Our study was relatively small (124 children with MAM enrolled) and the duration of intervention was short (3 months). Moreover, it was not designed to investigate stunting as a primary outcome^24,25^. It will be important to perform larger, longer controlled studies in Bangladesh, as well as in different geographic/sociodemographic settings, to (i) examine the generalizability of the effects of MDCF-2 on stunting without concurrent wasting, versus when both are present in the same child, and (ii) assess the impact of timing (i.e., chronologic age) of initiation of intervention based on the representation of target MAGs in childrens’ gut microbial communities - a process that could be guided by affordable point-of-care diagnostic approaches (e.g., qPCR assays of growth-discriminatory MAGs). In addition, it is important to consider and optimize the dose of the MDCF-2 formulation – especially its constituent glycans - and the duration of treatment. Moreover, the background diet consumed by children may have a significant impact on their growth; based on data from food frequency questionnaires, we had previously found that children who consumed more of the complementary food types present in MDCF-2 in their normal diets during the intervention were more likely to be in the upper quartile of improved weight gain (WLZ)^25^.

Taken together, our findings suggest that the effects of MDCF-2 on the gut microbiota of undernourished Bangladeshi children extend beyond weight gain to include improvements in linear growth. Dissecting the mechanistic underpinnings of the effects of the microbial community on growth requires additional research. For example, preclinical studies conducted in gnotobiotic mice colonized with defined communities of cultured, genome-sequenced bacteria corresponding to WLZ- and/or LAZ-associated MAGs represent one approach for dissecting the contributions of MDCF-2 responsive organisms to host pathways involved in ponderal and linear growth as well as other aspects of development, including neurodevelopment. A precedent for such an approach has been established. By combining analyses of microbial community assembly, microbial gene expression and metabolism of glycan constituents of MDCF-2 with single nucleus RNA-Seq and mass spectrometric analyses of the intestine we have confirmed the key role played by specific *P. copri* strains in metabolizing MDCF-2 glycans, promoting weight gain, and stimulating the activities of metabolic pathways involved in lipid, amino acid, carbohydrate plus other facets of energy metabolism in intestinal epithelial cells^53^. A similar strategy should now be employed to examine the role of cultured representatives of the LAZ-associated MAGs identified in the current study on different aspects of linear growth, including accretion of lean body mass, bone development, and maturation of the epiphyseal plates.

## Data Availability

The data supporting the findings of this study are available within the supplementary materials; shotgun sequencing data in raw format (prior to post-processing and data analysis) have been deposited at the European Nucleotide Archive (ENA) under study accession PRJEB66487.

https://www.ebi.ac.uk/ena/browser/view/PRJEB66487

## Contributors

I.M., with assistance from H.H.R., M.M., T.H. conducted the long-term follow-up for the clinical trial and collected anthropometry and clinical metadata. I.M., M.C.H. and M.J.B. analyzed dietary data and anthropometry as reported in Figure 1 and Tables 1 and 2. M.C.H. analyzed the enteropathogen qPCR data, mapped shotgun sequencing data to MAGs and conducted analyses of MAG abundance associations with prior MDCF-2 treatment and/or anthropometry that are reported in Figure 2. S.J.H. analyzed the correlations between plasma protein levels and LAZ or WLZ scores reported in Figure 3. M.J.B. oversaw databases of clinical metadata and the biospecimen archive. T.A. and J.I.G. supervised this research project. I.M., M.J.B. M.C.H. and J.I.G. wrote the paper with invaluable input from co-authors.

## Acknowledgments

We are grateful to the families of study participants and icddr,b investigators and staff for their contributions to the collection of biospecimens and data reported here. We appreciate the support of the Bill and Melinda Gates Foundation and the Governments of Bangladesh and Canada for providing core/unrestricted support to icddr,b. We thank Su Deng, Kazi Ahsan, Martin Meier, Jessica Hoisington-Lopez, and MariaLynn Crosby for their technical assistance with processing biospecimens. The support of members of the Genome Technology Access Core (GTAC) at Washington University School of Medicine in shotgun sequencing of fecal DNA and enteropathogen qPCR is greatly appreciated.

## List of Supplementary Materials

**Supplementary Table S1.** Analyses of anthropometry data

**Supplementary Table S2:** Analysis of enteropathogen abundances

**Supplementary Table S3:** Analysis of metagenome-assembled genome (MAG) abundances

**Supplementary Table S4.** Plasma protein levels related to anthropometric responses and treatment

